# Penicillin Allergy as an Instrumental Variable for Estimating Antibiotic Effects on Resistance

**DOI:** 10.1101/2024.08.19.24312204

**Authors:** Yaki Saciuk, Daniel Nevo, Michal Chowers, Uri Obolski

**Affiliations:** Department of Epidemiology and Preventive Medicine, School of Public Health, Faculty of Medical & Health Sciences Tel Aviv University, Tel Aviv, Israel; Kahn Sagol Maccabi (KSM) Research & Innovation Center, Maccabi Healthcare Services, Tel Aviv, Israel; Department of Statistics and Operations Research, Faculty of Exact Sciences, Tel Aviv University, Tel Aviv, Israel; Faculty of Medical & Health Sciences, Tel Aviv University, Tel Aviv, Israel; Meir Medical Center, Kfar Saba, Israel

## Abstract

Antibiotic resistance is influenced by prior antibiotic use, but precise causal estimates are limited. This study uses penicillin allergy as an instrumental variable (IV) to estimate the causal effect of antibiotics on resistance. A retrospective cohort of 36,351 individuals with *E. coli* positive urine cultures and prior outpatient antibiotic use, with outcomes assessed up to one year post-exposure, was analyzed using data from Maccabi Healthcare Services (MHS), the second-largest non-profit health fund in Israel. IV methods estimated risk differences (RD) and numbers needed to harm (NNH) for penicillin versus other antibiotics. The RD for resistance was 11.4% (95% CI: 7.6%, 15.4%) for amoxicillin/clavulanic acid, 14.1% (95% CI: 9.0%, 19.4%) for ampicillin, and 0.8% (95% CI: 0.2%, 1.4%) for piperacillin/tazobactam, with NNHs of 8.8, 7.1, and 122.0, respectively. Risks declined over time since exposure. Gentamicin, used as a negative control, showed no effect (95% CI: -2.4%, 1.8%). When directly comparing penicillin and quinolone effects on their respective AMR, penicillin use within 180 days increased resistance to amoxicillin/clavulanic acid by an RD of 17.8% (95% CI: 2.1%, 35.2%; NNH: 5.6), while quinolones raised ciprofloxacin resistance by 43.7% (95% CI: 29.9%, 59.4%; NNH: 2.3). These findings provide quantitative evidence of the impact of prior penicillin use on resistance, with implications for clinical practice and prescription policies.

## Introduction

Antibiotics are vital in combating bacterial infections, yet their effectiveness is diminishing due to increased antimicrobial resistance (AMR), partly stemming from overuse and misuse ^1,2^. The susceptibility of an infection to antibiotics is typically determined through microbiological testing, a process that can take several days. Consequently, a common scenario contributing to antibiotic misuse arises when patients are treated with antibiotics before the causative pathogen and its AMR profile are identified—an approach known as empiric therapy. Once antibiotic susceptibility results are available, they are reported as minimum inhibitory concentrations (MICs)—the minimum concentration of an antibiotic required to inhibit the growth of the pathogen—or categorized into susceptible, intermediate, or resistant groups based on established MIC thresholds^3^. To enhance responsible prescription of antibiotics, guidelines have been developed that facilitate treatment decisions based on anticipated pathogens, resistance profiles, and related healthcare costs of different antibiotics^3^.

AMR arises from both individual and population-level antibiotic use ^4^. At the population-level, the overall volume and patterns of antibiotic play a significant role in driving resistance, as resistant pathogens spread between individuals ^4,5^. At the individual level, antibiotic use selects for infection and colonization with resistant bacterial strains ^5,6^. In recent years, there has been a growing focus on the potential of different antibiotics to induce future resistance on an individual-level. For example, quinolones, previously the primary choice for urinary tract infections and community-acquired pneumonia, are now often disfavored due to their high potential for resistance induction^7^.

Nonetheless, a significant gap in current research is the lack of quantitative, causal assessments of the risk of future resistance caused by different antibiotics. Randomized control trials, the benchmark for causal effect estimation, present logistical difficulties in this context, including limited patient follow up and the requirement for numerous studies to address various clinical scenarios and antibiotic combinations. Consequently, ranking of antibiotics by their collateral resistance cost is often reached by expert opinion ^8–10^. Alternatively, cohort studies aimed at evaluating the effects of antibiotics are susceptible to numerous confounding mechanisms that undermine the causal validity of their results^5,11^.

Potential confounders, such as the indication for antibiotic use, illness severity at presentation, and patient demographics, present substantial challenges for retrospective studies in this field; researchers often struggle to access these variables. However, advances in causal inference theory have substantially improved our ability to discern causal effects from observational data. In particular, the use of instrumental variables (IVs) can provide reliable estimates of causal effects, when the appropriate methodology and falsification tests are applied ^12,13^, emulating natural-experiment-like conditions.

Here, we propose to employ reported allergy to penicillin as such an IV. Reported penicillin allergy satisfies the essential conditions needed for a valid IV analysis. First, it causally influences prescription practices^14^, leading physicians to opt for non-penicillin antibiotics upon presentation of penicillin allergy. Second, reported penicillin allergy also inherently does not directly affect AMR of bacterial populations.

By controlling for measured covariates which have the potential to confound the relationship between the IV (allergy to penicillin) and the outcome (AMR), we can further bolster the assumption of the causal nature of this association. Moreover, the setting of antibiotic susceptibility testing provides a falsification test for this assumption. We can utilize resistance to antibiotics outside the penicillin group as a negative control outcome (NCO) ^13,15,16^, which could offer additional support for the validity of this assumption. The essential causal relationships assumed in our study are concisely depicted in the directed acyclic graph (DAG)^17^ in Figure 1.

**Figure 1:**
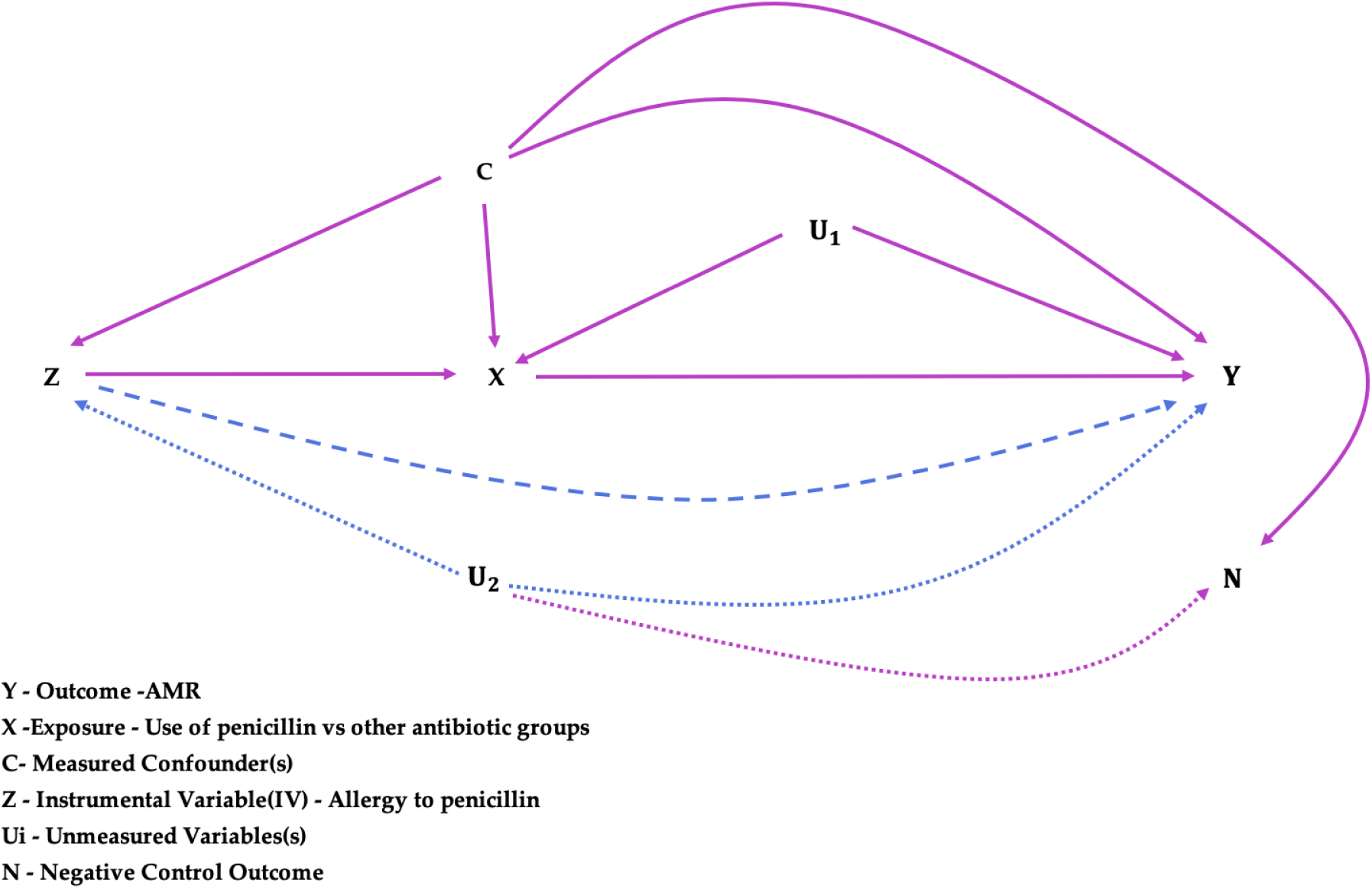
Causal relations in the instrumental variable (IV) framework can be depicted using Directed Acyclic Graphs (DAGs). The interest lies in determining the effect of the exposure (*X*) on the outcome (*Y*). This effect may not be identifiable—i.e., causally estimable—in the presence of unmeasured confounders (*U_1_*). However, employing a valid IV (*Z*) may enable the identification of this effect. First, *Z* must influence the exposure *X*, a condition known as the Relevance condition. Furthermore, *Z* should affect *Y* solely through *X*—a condition termed Exclusion Restriction. Measured confounders (*C*) and unmeasured variables (*U*) may impact *Z*, *X* and *Y*, potentially confounding their association. Adjusting for *C* blocks the *Z* <-*C* -> *Y* path, strengthening the third requirement—the Independence condition. However, the presence of an unmeasured variable (*U_2_*) could still compromise this condition and hinder the identification of the causal effect of *X* on *Y*. Dashed and dotted lines represent violations of the Exclusion Restriction and Independence conditions, respectively. A negative control outcome (*N*) should share a similar set of measured common causes (*C*) with *Y* and *Z*. If the unobserved common causes of *Z* and *Y* overlap with those of *Z* and *N* (*U_2_*), then *N* is considered *U-comparable* to *Y*. Hence, if the independence condition is met, we would naturally expect *Z* to not be associated with *N*, when adjusting for *C*.

In this study, we utilize a large and comprehensive medical records database and leverage IV analysis to estimate the effects of consumption of penicillins, compared to other antibiotics, on AMR in subsequent bacterial cultures. Specifically, we estimate the Risk Difference (RD) and Number Needed to Harm (NNH) of AMR in *E. coli.* We further estimate the effects of antibiotic consumption on AMR over different time periods following antibiotic use. To do this, we adjust for multiple variables to eliminate potential IV-outcome confounding and apply inverse probability of censoring weighting (IPCW) to mitigate selection bias. Finally, we perform negative control analyses to ascertain the validity of our IV.

## Results

Within the primary research population, encompassing 1,469,624 individuals who received at least one outpatient antibiotic prescription from January 2013 to December 2022, a subset of 36,351 individuals underwent at least one AMR test panel within a following 12-month period. In total, 41,647 AMR test panels were administered. The distribution of patient characteristics and selection criteria for the research population are further illustrated in Figure 2, with detailed patient characteristics provided in Table 1. To verify the IV Relevance condition, we computed the first-stage adjusted OLS regression, which yielded significant Kleibergen-Paap F-statistics and IV coefficients of approximately 23%, indicating a strong link between the IV and the exposure (Table S1).

**Figure 2:**
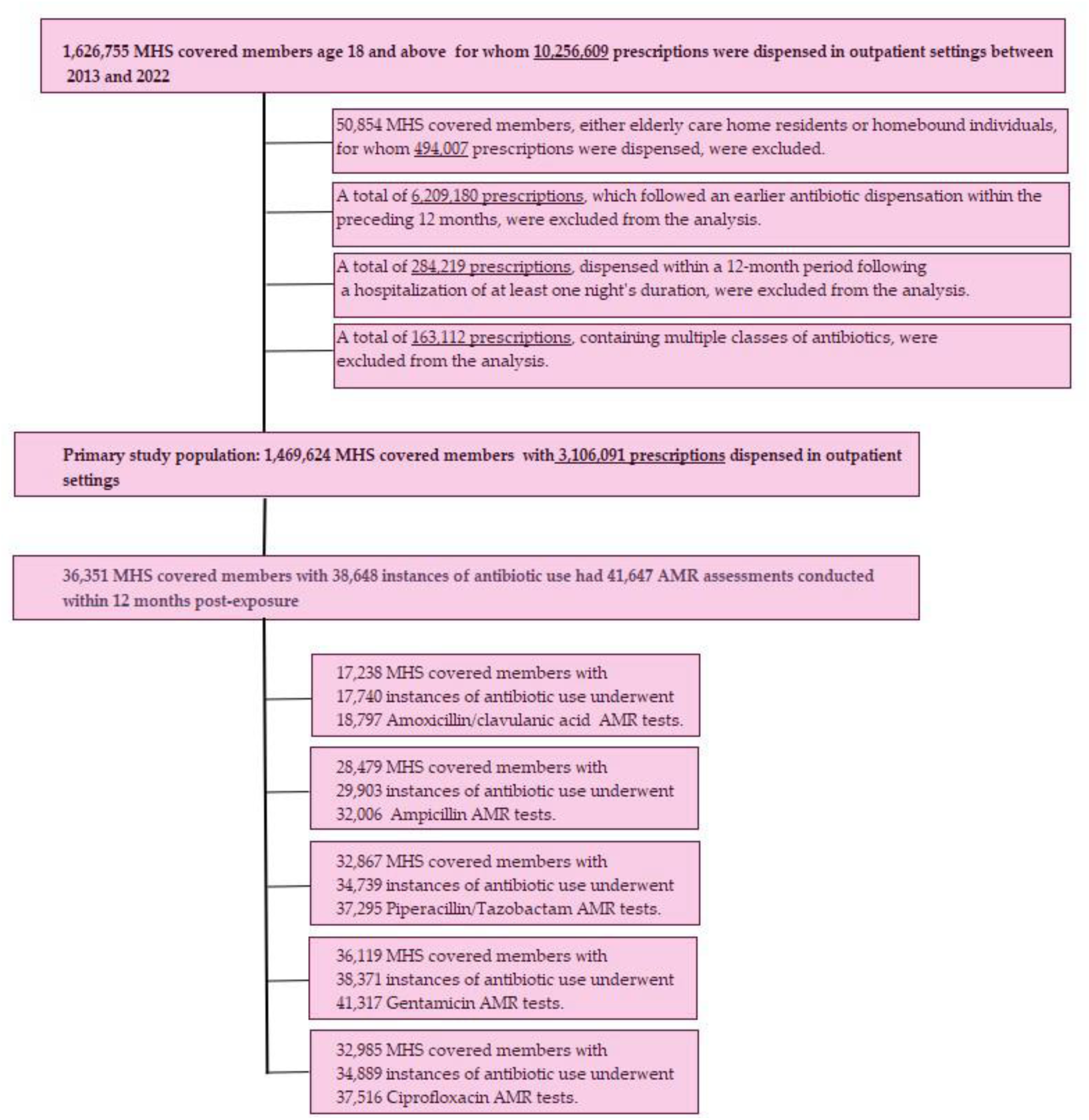
Study population flow chart.

**Table 1:**
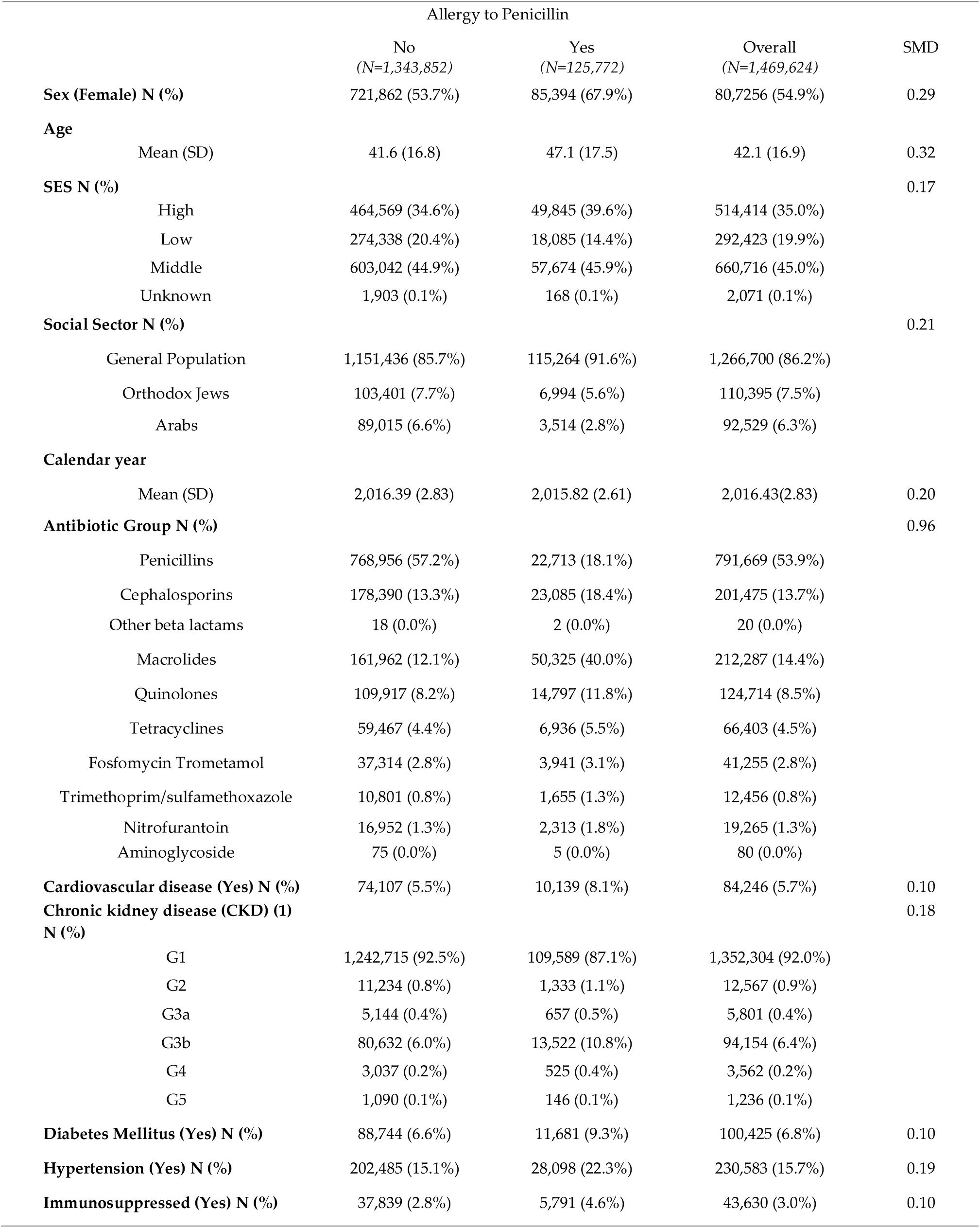
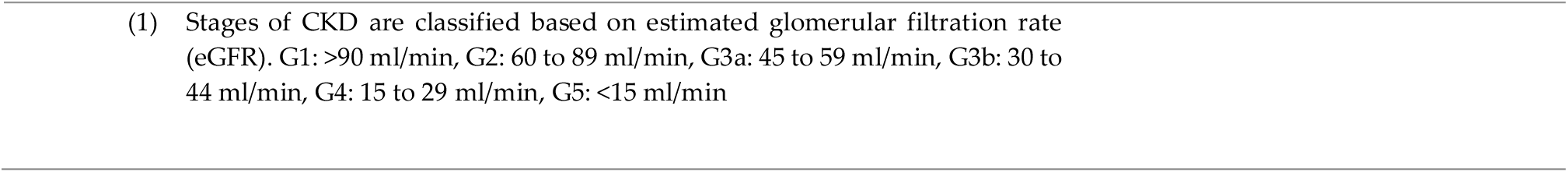
Characteristics of the Primary Research Population at First Eligible Antibiotic Use (2013-2022), stratified by reported Penicillin Allergy.

| Allergy to Penicillin |  |  |  |  |
| --- | --- | --- | --- | --- |
|  | No<br>(N=1,343,852) | Yes<br>(N=125,772) | Overall<br>(N=1,469,624) | SMD |
| **Sex (Female) N (%)** | 721,862 (53.7%) | 85,394 (67.9%) | 80,7256 (54.9%) | 0.29 |
| **Age** |  |  |  |  |
| Mean (SD) | 41.6 (16.8) | 47.1 (17.5) | 42.1 (16.9) | 0.32 |
| **SES N (%)** |  |  |  | 0.17 |
| High | 464,569 (34.6%) | 49,845 (39.6%) | 514,414 (35.0%) |  |
| Low | 274,338 (20.4%) | 18,085 (14.4%) | 292,423 (19.9%) |  |
| Middle | 603,042 (44.9%) | 57,674 (45.9%) | 660,716 (45.0%) |  |
| Unknown | 1,903 (0.1%) | 168 (0.1%) | 2,071 (0.1%) |  |
| **Social Sector N (%)** |  |  |  | 0.21 |
| General Population | 1,151,436 (85.7%) | 115,264 (91.6%) | 1,266,700 (86.2%) |  |
| Orthodox Jews | 103,401 (7.7%) | 6,994 (5.6%) | 110,395 (7.5%) |  |
| Arabs | 89,015 (6.6%) | 3,514 (2.8%) | 92,529 (6.3%) |  |
| **Calendar year** |  |  |  |  |
| Mean (SD) | 2,016.39 (2.83) | 2,015.82 (2.61) | 2,016.43(2.83) | 0.20 |
| **Antibiotic Group N (%)** |  |  |  | 0.96 |
| Penicillins | 768,956 (57.2%) | 22,713 (18.1%) | 791,669 (53.9%) |  |
| Cephalosporins | 178,390 (13.3%) | 23,085 (18.4%) | 201,475 (13.7%) |  |
| Other beta lactams | 18 (0.0%) | 2 (0.0%) | 20 (0.0%) |  |
| Macrolides | 161,962 (12.1%) | 50,325 (40.0%) | 212,287 (14.4%) |  |
| Quinolones | 109,917 (8.2%) | 14,797 (11.8%) | 124,714 (8.5%) |  |
| Tetracyclines | 59,467 (4.4%) | 6,936 (5.5%) | 66,403 (4.5%) |  |
| Fosfomycin Trometamol | 37,314 (2.8%) | 3,941 (3.1%) | 41,255 (2.8%) |  |
| Trimethoprim/sulfamethoxazole | 10,801 (0.8%) | 1,655 (1.3%) | 12,456 (0.8%) |  |
| Nitrofurantoin | 16,952 (1.3%) | 2,313 (1.8%) | 19,265 (1.3%) |  |
| Aminoglycoside | 75 (0.0%) | 5 (0.0%) | 80 (0.0%) |  |
| **Cardiovascular disease (Yes) N (%)** | 74,107 (5.5%) | 10,139 (8.1%) | 84,246 (5.7%) | 0.10 |
| **Chronic kidney disease (CKD) (1) N (%)** |  |  |  | 0.18 |
| G1 | 1,242,715 (92.5%) | 109,589 (87.1%) | 1,352,304 (92.0%) |  |
| G2 | 11,234 (0.8%) | 1,333 (1.1%) | 12,567 (0.9%) |  |
| G3a | 5,144 (0.4%) | 657 (0.5%) | 5,801 (0.4%) |  |
| G3b | 80,632 (6.0%) | 13,522 (10.8%) | 94,154 (6.4%) |  |
| G4 | 3,037 (0.2%) | 525 (0.4%) | 3,562 (0.2%) |  |
| G5 | 1,090 (0.1%) | 146 (0.1%) | 1,236 (0.1%) |  |
| **Diabetes Mellitus (Yes) N (%)** | 88,744 (6.6%) | 11,681 (9.3%) | 100,425 (6.8%) | 0.10 |
| **Hypertension (Yes) N (%)** | 202,485 (15.1%) | 28,098 (22.3%) | 230,583 (15.7%) | 0.19 |
| **Immunosuppressed (Yes) N (%)** | 37,839 (2.8%) | 5,791 (4.6%) | 43,630 (3.0%) | 0.10 |

- - (1) Stages of CKD are classified based on estimated glomerular filtration rate (eGFR). G1: >90 ml/min, G2: 60 to 89 ml/min, G3a: 45 to 59 ml/min, G3b: 30 to 44 ml/min, G4: 15 to 29 ml/min, G5: <15 ml/min -

We analyzed varying post-exposure follow-up times for two main reasons. First, the impact of antibiotic exposure on AMR diminishes over time as the perturbation to the patient’s microbiome subsides, with most associations likely decaying after a year^6,18^, making estimates at different time points valuable. Second, identifying this time-dependent effect (or its absence, in the NCO) further validates our analyses. In the initial up to 90 days post-exposure, penicillin users exhibited significantly higher RDs for AMR within the penicillin group compared to other treatments. Specifically, resistance to amoxicillin/clavulanic acid, ampicillin, and piperacillin/tazobactam exhibited RDs of 19.0% (95% CI: 11.8%, 26.1%), 22.6% (95% CI: 14.6%, 31.0%), and 2.5% (95% CI: 1.4%, 3.5%), respectively. The corresponding NNH values were 5.3, 4.4, and 40.2 (Table 2). These RDs consistently remained statistically significant across various post-exposure outcome time points and exhibited a temporal decline, aligning with expected AMR reduction as time elapsed. In contrast, gentamicin’s RD, serving as a negative control, was close to zero and remained statistically non-significant at all time points and did not show a decreasing trend in the RD point estimate (Figure 3). It is worth emphasizing that the estimate within each interval encapsulates a weighted average of the RD over the entire period, rather than an RD estimated at a specific time point.

**Figure 3:**
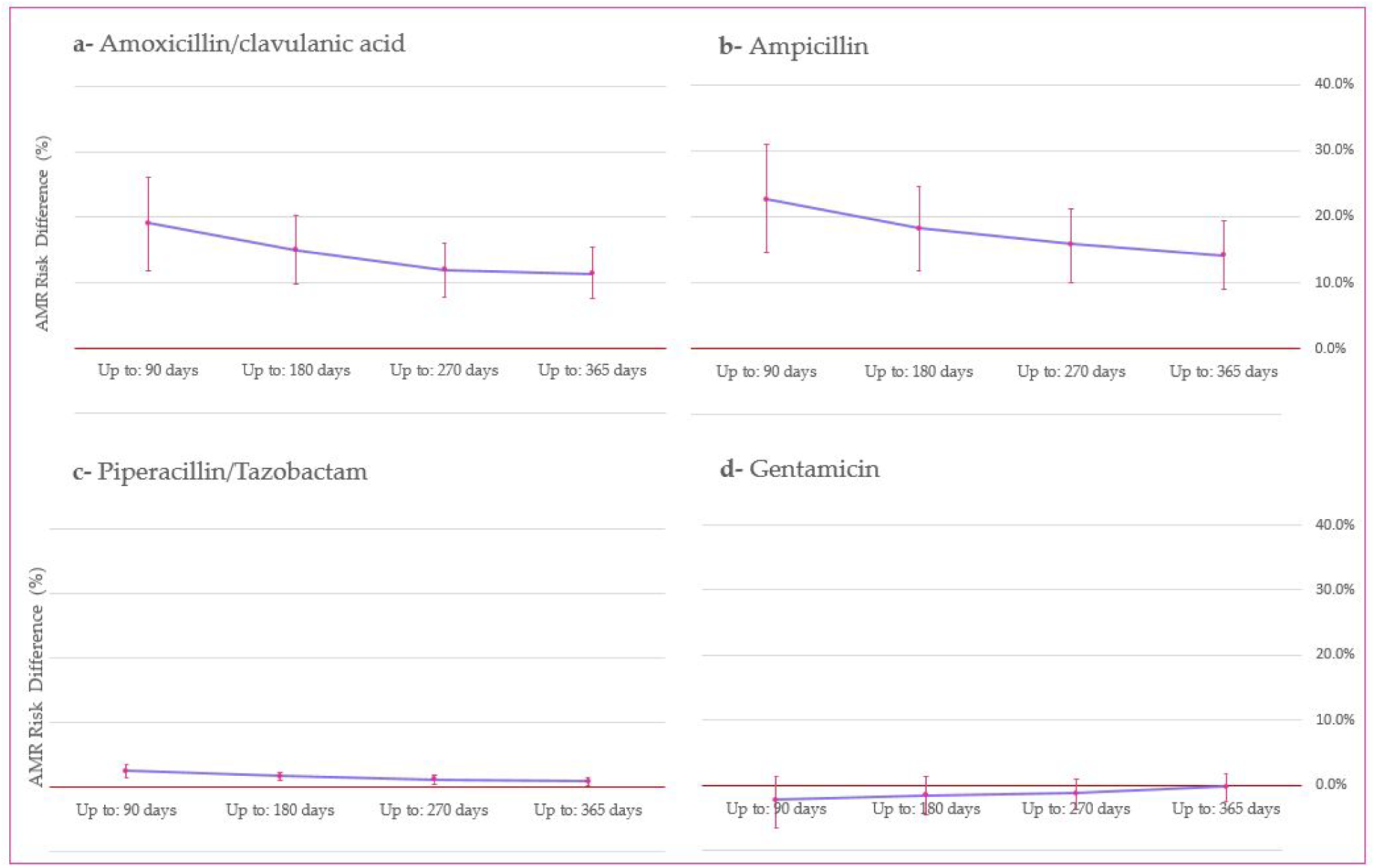
Instrumental Variable (IV)-Estimated Risk Differences for Antimicrobial Resistance (AMR) Over Time Post-Exposure to Penicillins Compared to Other Antibiotics. AMR outcomes include: (a) amoxicillin/clavulanic acid (n=18,797 tests), (b) ampicillin (n=32,006 tests), (c) piperacillin/tazobactam (n=37,295 tests), and (d) gentamicin (n=41,317 tests). Bars represent 95% confidence intervals (CIs) around the mean risk differences, derived from bootstrap sampling (3,000 iterations). The x-axis displays the number of days following exposure to penicillins. Risk differences compare resistance outcomes following exposure to penicillins versus other antibiotics. Detailed numerical values, including risk differences and corresponding confidence intervals, are provided in Table 2

**Table 2:** Instrumental Variable (IV)-Estimated Risk Differences of Antimicrobial Resistance (AMR) Rates: A Comparison between Exposure to Penicillins and Other Antibiotics Across <u>Varying Time</u> Points Post-Exposure.

| AMR Antibiotic | Follow up length | Number of AMR Tests | RD IV (1)(2) | 95% C.I. (1)(2) |  | NNH |
| --- | --- | --- | --- | --- | --- | --- |
|  |  |  |  | Lower Bound (2.5%) | Upper Bound (97.5%) |  |
| Amoxicillin/clavulanic acid | Up to: 90 days | 7,502 | 19.0% | 11.8% | 26.1% | 5.3 |
|  | Up to: 180 days | 12,393 | 14.9% | 9.9% | 20.2% | 6.7 |
|  | Up to: 270 days | 16,027 | 11.9% | 7.8% | 16.1% | 8.4 |
|  | Up to: 365 days | 18,797 | 11.4% | 7.6% | 15.4% | 8.8 |
| Ampicillin | Up to: 90 days | 13,776 | 22.6% | 14.6% | 31.0% | 4.4 |
|  | Up to: 180 days | 21,828 | 18.2% | 11.9% | 24.6% | 5.5 |
|  | Up to: 270 days | 27,587 | 15.9% | 10.1% | 21.2% | 6.3 |
|  | Up to: 365 days | 32,006 | 14.1% | 9.0% | 19.4% | 7.1 |
| Piperacillin/Tazobactam | Up to: 90 days | 15,203 | 2.5% | 1.4% | 3.5% | 40.2 |
|  | Up to: 180 days | 24,639 | 1.6% | 0.9% | 2.2% | 61.7 |
|  | Up to: 270 days | 31,730 | 1.1% | 0.4% | 1.7% | 87.7 |
|  | Up to: 365 days | 37,295 | 0.8% | 0.2% | 1.4% | 122.0 |
| Gentamicin (Negative Control) | Up to: 90 days | 17346 | -2.3% | -6.5% | 1.4% | -43.5* |
|  | Up to: 180 days | 27824 | -1.5% | -4.5% | 1.3% | -66.2* |
|  | Up to: 270 days | 35447 | -1.3% | -3.8% | 0.8% | -79.4* |
|  | Up to: 365 days | 41317 | -0.2% | -2.4% | 1.8% | -500.0* |

(1) Weighted with inverse probability of censoring weights (IPCW)
(2) Adjusted for sex, age, ses, social sector, comorbidities (CKD, cardio-vascular disease, diabetes mellitus, hypertension and immunosuppression), calendar time, previous urine culture tests count, and pervious antibiotic uses count
\* The negative NNH is interpreted as the absolute value of the NNH when switching the penicillins to be the comparator group.

We conducted several sensitivity analyses to ascertain the robustness of our results. First, the presented results incorporate IPCW adjustments, and unweighted analyses, exhibiting similar results, are available in Table S2 for comparison. Second, although adjusted for in the primary analysis, the socio-demographic differences between penicillin-allergic and non-allergic groups could potentially affect the validity of the results. To address this concern, we conducted sensitivity analyses using full matching ^19,20^ on covariates and replaced the standard linear model with a probit model in the first stage of the 2SLS procedure^21^. The results again largely corroborated our original findings (Table S3).

Finally, to address the composition of the penicillin group, we repeated our original analysis while limiting the exposure to either amoxicillin/clavulanic acid, amoxicillin, or amoxicillin with other penicillins. The consistent results from this analysis are displayed in Table S4.

We then directly compared the effects of penicillin and quinolone exposure on AMR, focusing on a 180-day follow-up period. This follow-up duration was chosen to balance the trade-off between effect size and sample size, as the subgroup sizes became smaller for longer follow-up periods. To evaluate quinolone resistance, we analyzed susceptibility results for ciprofloxacin, a commonly tested-for-resistance quinolone within Maccabi Healthcare Services (MHS). The findings, summarized in Table 3 and Figure 4, indicate prior penicillin use increases resistance, relative to prior fluoroquinolone use in three penicillin antibiotics, albeit the ampicillin resistance outcome was not statistically significant. Conversely, prior quinolone exposure, relative to penicillin exposure, presented a pronounced influence on subsequent quinolone resistance (with a negative RD due to quinolones being the control group). The RD for amoxicillin/clavulanic acid was 17.8% (95% CI: 2.1%, 35.2%), for ampicillin 8.0% (95% CI: -5.3%, 22.6%), for piperacillin/tazobactam 2.8% (95% CI: 0.9%, 4.6%), for gentamicin -5.4% (95% CI: -14.6%, 2.8%), and for ciprofloxacin -43.9% (95% CI: -59.4%, -29.9%).

**Figure 4:**
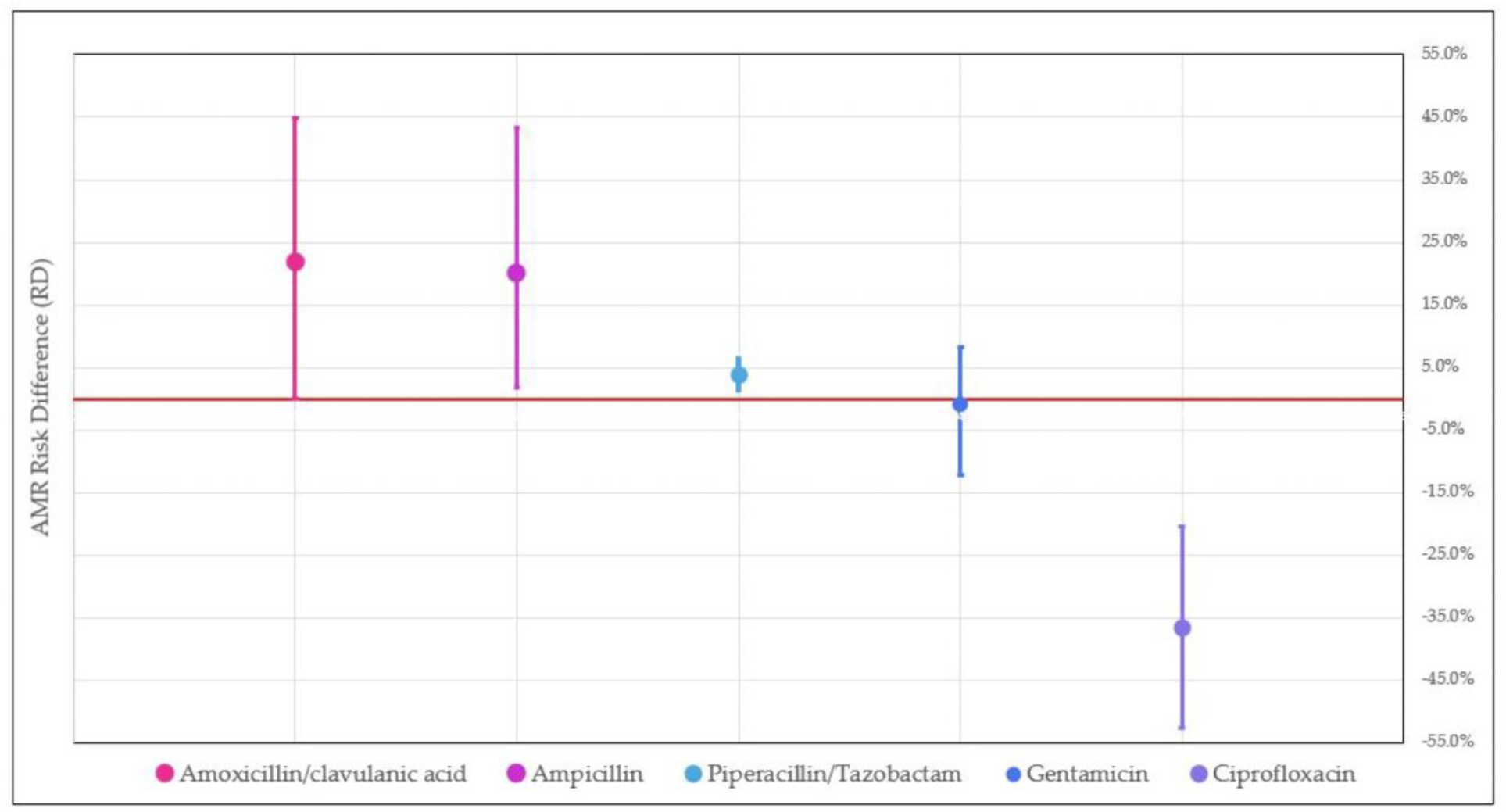
Instrumental Variable (IV)-Estimated Risk Differences for Antimicrobial Resistance (AMR) Between Penicillins and Quinolones Within 180 Days After Exposure. Bars represent 95% confidence intervals (CIs) derived from bootstrap sampling (3,000 iterations. The x-axis displays the tested AMR outcomes, which include: amoxicillin/clavulanic acid (n=4,918 tests), ampicillin (n=10,003 tests), piperacillin/tazobactam (n=10,564 tests), gentamicin (n=12,291 tests), and ciprofloxacin (n=11,073 tests). Risk differences compare resistance outcomes following exposure to penicillins versus quinolones. Detailed numerical values, including risk differences and corresponding confidence intervals, are provided in Table 3.

**Table 3:**
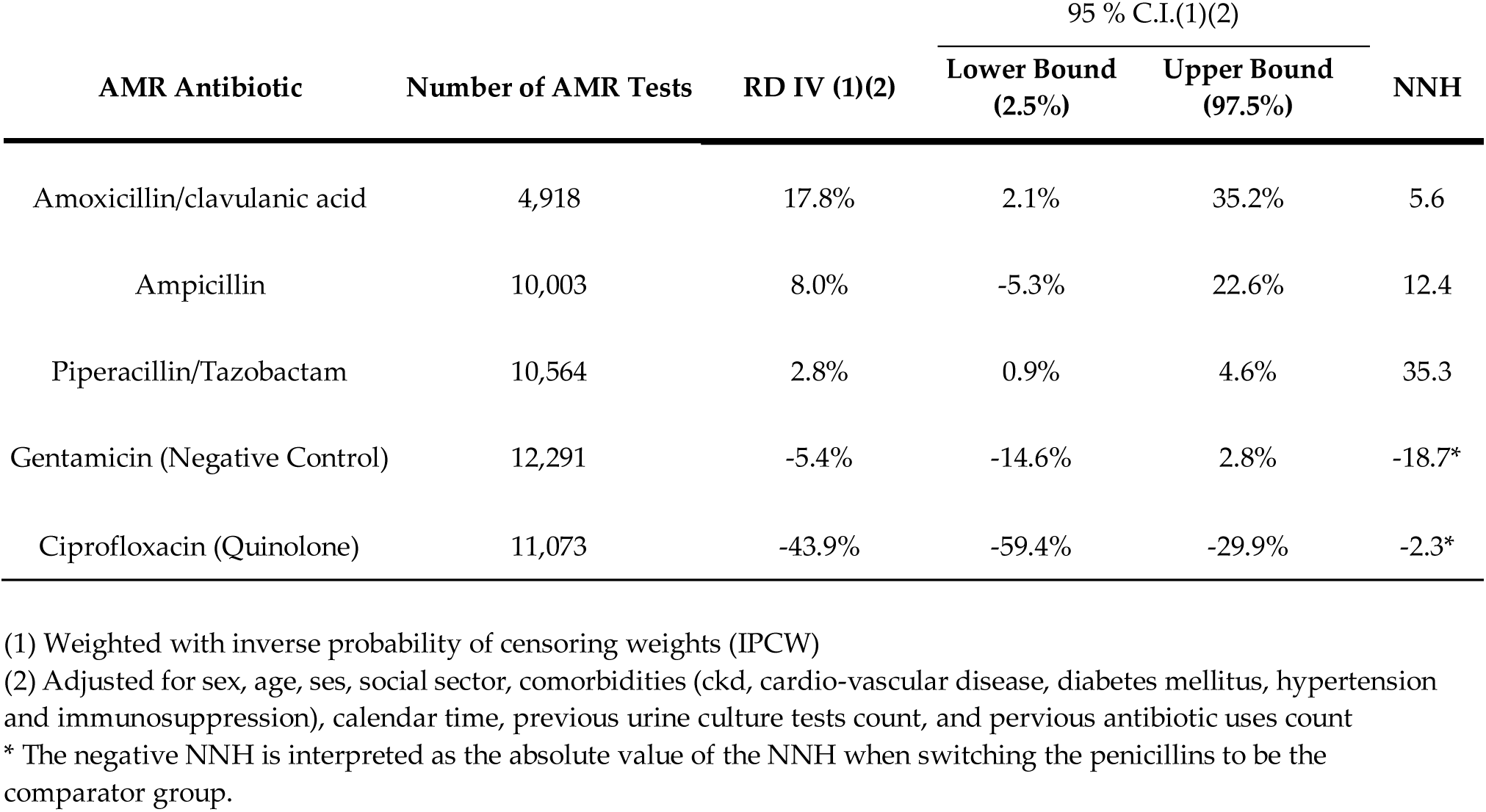
Instrumental Variable (IV)-Estimated Risk Differences of Antimicrobial Resistance (AMR): Direct Comparison of Penicillin vs. Quinolones Within 180 Days Post-Exposure.

We conducted pairwise comparisons between penicillin and each antibiotic group within the control arm, excluding quinolones, aminoglycosides, and other beta-lactams, within the 180-day window post-exposure (Table S5). The RDs varied somewhat. Comparisons with cephalosporins, another beta-lactam, yielded statistically insignificant RDs for penicillins. Similarly, comparisons of penicillins versus other groups for gentamicin AMR also yielded statistically insignificant RDs.

## Discussion

This study exploits a unique phenomenon, namely purported penicillin allergy, to estimate the causal effects of prior antibiotic use on antibiotic resistance, in real-world primary care settings. Our analysis quantifies the impact of previous use of penicillins on resistance rates of future infections to the three penicillins. The largest effect was observed in resistance to ampicillin, followed by amoxicillin/clavulanic acid, and finally piperacillin/tazobactam. The low NNH values estimated for amoxicillin/clavulanic acid and ampicillin suggest that consumption of penicillins may frequently lead to AMR to these antibiotics upon infections. In contrast, the higher NNH for piperacillin/tazobactam indicates a comparatively lower cost of resistance, suggesting that only a small number of patients exposed to penicillins would lose the ability to be effectively treated with this antibiotic due to resistance.

In line with both biological reasoning and previous knowledge, all three outcomes demonstrated a gradual decrease in the RD for antibiotic resistance with each subsequent time point, reflecting the diminishing impact of prior antibiotic use ^18^. In contrast, gentamicin, the negative-control antibiotic, displayed no such effect nor trend, further affirming the validity of our analysis. In our additional analysis, we compared past usage of penicillin with that of quinolones on corresponding AMR tests within the 180-day post-exposure. The past use of penicillins was linked to resistance rates for penicillins; but this association was lower than the association between past use of quinolones, and higher resistance to ciprofloxacin. This finding is consistent with previous evidence that quinolones usage leads to substantial increase in resistance ^18,22–24^. However, the current study provides an unbiased, causally justified comparison between the antibiotic classes.

As with any study attempting to infer causality from observational data, this study has its limitations. The IV analysis performed here requires several specific assumptions. The Relevance condition has been confirmed through the coefficients obtained from the first-stage regression models. With respect to the Independence condition, Table 1 contrasts differences in demographic characteristics between individuals with and without a penicillin allergy. While the differences are not pronounced, they are present: the allergic population is predominantly female, older, and of higher socioeconomic status, suggesting a possible behavioral rather than biological basis for the reporting of penicillin allergy ^25^. The integrity of this condition was kept by adjusting for these variables. Moreover, the near-zero effect observed in the NCO, and the lack of temporal trend, further reassures that this condition is maintained.

To estimate the average treatment effect using IVs, we assume that treatment effects are constant across the population. If this assumption is violated, the estimated effect can still be interpreted if the Monotonicity assumption holds^12^. This assumption, also known as the *no defiers* assumption, means that no individual will consistently choose a treatment contrary to their IV assignment. In our study, it implies that individuals who would receive penicillin if not allergic would also avoid it if allergic. Under this assumption and additional statistical assumptions, the estimated effect is an average of Complier Average Causal Effects (CACEs) or Local Average Treatment Effects (LATEs)^12^. The CACE/LATE represents the impact on individuals who comply with the treatment indicated by the IV, known as *compliers*.

The study’s final limitation concerns the generalizability of its estimated effects. We used a comprehensive dataset that well-represents antimicrobial resistance of *E. coli* urine isolates in Israel. However, the study outcomes are likely influenced by the local variations in resistance patterns of pathogens during the period and location of the study (Figure S1). This is a common occurrence in infectious diseases, the dynamics of which are dictated by local biological and behavioral factors. Furthermore, our inclusion criteria, which excluded individuals who had purchased antibiotics or been hospitalized within the previous 12 months, may have resulted in a somewhat healthier-than-average sample, potentially limiting generalizability to other populations. Nevertheless, our study also provides researchers with a novel approach and detailed methodology to estimate the effects of antibiotic exposure on future resistance, to apply it to specific locations and time periods.

In conclusion, our findings have implications for both guidelines and clinical practice, and methodological approaches to the epidemiology of antibiotic resistance. Clinically, our results provide causal estimates of the additional risk and NNH impacted by prior use of penicillins and quinolones in the outpatient settings, over different periods since exposure. Such estimates should also play a substantial role in the cost-benefit considerations when determining empiric antibiotic treatment guidelines. Specifically, penicillins induce substantial future resistance to amoxicillin/clavulanic acid, ampicillin, and to a lower extent piperacillin tazobactam. Moreover, penicillins, compared to quinolones, appear to induce lower respective AMR, thereby enhancing their utility in subsequent treatments. Methodologically, we propose a novel approach, built on an original IV, that opens a venue for further research on antibiotic groups inducing allergic reactions, such as cephalosporins, fluoroquinolones, and sulfonamides.

## Methods

This research received IRB approval from Maccabi Healthcare Services (MHS) under protocol number MHS-0036-23.

### Data

Maccabi Healthcare Services (MHS), the second-largest non-profit health fund in Israel, covers 26.7% of the population (∼2.7 million in 2022) nationwide, with less than 1% disengagement. Membership in one of the four health funds is mandatory and non-excludable. MHS maintains a long-term centralized database of electronic medical records (EMRs), including extensive demographic and clinical data from both outpatients and hospitals.

### Study design

In this retrospective cohort study, data were collected on adult members (aged≥18 years) of MHS who purchased antibiotics in primary care between January 1, 2013, and December 31, 2022.

Individuals were eligible to contribute multiple entries to the study within the specified timeframe, provided they had not purchased antibiotics within 12 months prior to the entry date or been hospitalized for at least one night during the same period.

The exposed group consisted of all penicillin-group antibiotic purchases, while the control groups included purchases from the following antibiotic families: cephalosporins, macrolides, trimethoprim-sulfamethoxazole, quinolones, tetracyclines, fosfomycin, nitrofurantoin, aminoglycosides, and other beta-lactam antibiotics (Table 1).

The exposures to penicillins were comprised of 46% amoxicillin, 39% amoxicillin/clavulanic acid, and the remaining 15% of other penicillin derivatives. Although these antibiotics may serve to treat different indications, there is also substantial overlap between their targets and therefore physicians may sometimes prescribe them interchangeably^3^.

For both groups, the index date was set as the date of antibiotic purchase, and individuals were followed until either a positive urine culture test that prompted an AMR test (the outcome) or patient exclusion occurred. Censoring, in the sense of excluding the follow-up of patients to observe their outcomes, was applied in cases of subsequent antibiotic use, hospitalization, death, or reaching the conclusion of the follow-up period, which extended until 365 days after the index date or until December 31, 2022 - whichever came first. Potential selection bias due to such censoring was addressed using IPCW, as described below.

We excluded cases where more than one antibiotic family had been purchased to negate the influence of other antibiotic exposures. Furthermore, we excluded individuals residing in nursing homes and those who were homebound to mitigate potential confounding attributable to healthcare-associated infections or distinct prescription practices prevalent outside of primary care settings. This study design permitted the inclusion of multiple AMR tests for a single entry, if the specified exclusion and censoring criteria were fulfilled.

### Procedures

Individual-level demographic data for the study population included sex, age at the index date, socio-economic status, and social sector indication. The socioeconomic status of residential areas was assessed using coded Geographical Statistical Areas (GSAs), the smallest units recognized by the Israeli census. Assigned by Israel’s Central Bureau of Statistics, these GSAs are rated on a scale from 1 (lowest) to 10, based on multiple parameters. Data on chronic conditions were obtained from MHS’ automated registries, covering cardiovascular diseases ^26^, hypertension ^27^, diabetes^28^, chronic kidney disease (CKD)^29^, and immunocompromised states. We identified penicillin allergy using two complementary sources: ICD-9 diagnosis codes (V14.0) and physician-documented mentions of allergy in the free-text sections of medical records. While free-text documentation was available only for cases already flagged with ICD-9 codes, no instances were observed where a free-text note indicated a penicillin allergy without a corresponding diagnosis code. Importantly, the observed prevalence of penicillin allergy in our cohort (8.5%) aligns well with the 8–12% prevalence reported in the literature^25,30,31^, supporting the validity of our identification methods.

In Israel, the procurement of antibiotics is strictly regulated, requiring a prescription for purchase and all transactions related to the acquisition of antibiotics by members of MHS to be systematically recorded and digitized within the EMR system.

### Outcomes

Our outcomes were the binary results of antibiotic susceptibility tests, with resistance coded as 1, at various follow-up times (90, 180, 270, and 365 days). We considered antibiotic resistance results within the penicillin group, comprising ampicillin, amoxicillin clavulanic acid and piperacillin/tazobactam, as well as ciprofloxacin within the fluoroquinolones group. We note that piperacillin/tazobactam was added as a penicillin-group outcome but not exposure, because susceptibility to it is routinely tested, while it is very rarely prescribed in the community settings, in Israel. Gentamicin, an aminoglycoside antibiotic, served as an NCO for the IV independence assumption ^15,16^(explained below). We chose gentamicin due to its lack of association with the penicillin/beta-lactam family, its relative frequency in AMR test panels, and its infrequent use in outpatient settings. The bacterial identification and antibiotic susceptibility testing were conducted using the automated VITEK 2 system, following Clinical and Laboratory Standards Institute guidelines. Intermediate results were considered as resistant.

Our analysis focused exclusively on *Escherichia coli* isolates, which accounted for 71.6% of the total samples. Utilizing IV analysis, we estimated the risk difference of antibiotic resistance between individuals exposed to penicillins and those exposed to alternative antibiotic groups at the various post-antibiotic use follow-up times.

### IV framework

We assert that antibiotic allergies, particularly penicillin^25^, can serve as an IV in studying antibiotic use and future AMR. Although often self-reported, misdiagnosed, or resolved^32^, perceived allergies influence physicians’ treatment choices, meeting the conditions for a valid IV.

To be valid, IVs need to fulfill three essential conditions ^12^. The first, known as the Relevance condition, stipulates that the IV must be associated with exposure. The second, the Exclusion Restriction condition, asserts that the IV’s impact on the outcome occurs exclusively through exposure, and not via alternative paths. The third, referred to as the Independence condition, requires that the association between the IV and the outcome is not confounded by external factors.

Provided that the linear model specification employed for estimation is valid, and under the assumption of a homogeneous treatment effect across the population, these conditions enable the identification and consistent estimation of the average treatment effect (ATE)^33^.

### Statistical Methods

We estimated the RD using Two Stage least squares (2SLS) regression^33–35^. To uphold the Independence condition, the estimation was adjusted for potential confounders of the IV (allergy to penicillin) and the outcome (AMR) association, including sex, age, socio-economic status, social sector, calendar time, and comorbidities such as diabetes mellitus (DM), chronic kidney disease (CKD), immunosuppression, hypertension and cardiovascular diseases. To address the potential influence of healthcare-seeking behavior on both penicillin allergy reporting and AMR, we used proxies like the count of previous antibiotic purchases and urine culture tests, adjusting for these variables in the model. All continuous variables were modeled using restricted cubic splines with five knots^36^. Potential confounders were measured at baseline. A DAG depicting the assumed causal relationships between penicillin allergy, AMR, and other relevant covariates is presented in Figure S2. Model diagnostics showed adequate fit, with all instrument coefficients and fitted values within the expected range of [0,1]. To evaluate the relevance condition, we used a *first-stage* ordinary least squares (OLS) regression, modeling the exposure as a function of the IV and confounding covariates. The condition’s validity was confirmed by examining the IV coefficient and Kleibergen-Paap F-statistics. The exposure variable, indicating the purchase of a penicillin antibiotic, was coded as binary. To maintain consistent directionality, the IV for penicillin allergy was coded as ’1’ for non-allergic and ’0’ for allergic individuals.

Finally, selection bias may be introduced into the study as not all individuals are subjected to resistance testing after a positive urine culture, and those who do may not be provided with relevant antibiotic panels. To address this potential bias and account for interdependent observations, we calculated 95% confidence intervals (CIs) using bootstrap resampling at the individual level, with sampling weights proportionally applied to the IPCW^37^. IPCW compensates for censored patients by giving more weight to similar non-censored patients, thus increasing their representation. The weights were obtained from a logistic regression model that estimated the likelihood of undergoing AMR testing. The model was adjusted for sex, age, socio-economic status, social sector, antibiotic group, calendar time, and comorbidities (CKD, immunosuppression, and cardiovascular diseases). Censoring probabilities and their distribution are in Table S6. Due to potential temporal variations in IPCW weights, which were time-dependent and varied across entries for individuals with multiple contributions, we used the average of these weights at the individual level in the bootstrap procedure.

Data were extracted from the MHS database using Microsoft SQL Server 14. All statistical analyses were performed in R version 3.6.2, and the ivtools package was used to estimate the instrumental variable (IV) models.

## Data Availability

According to the Israel Ministry of Health regulations, individual-level data cannot be shared openly. Specific requests for remote access to de-identified community-level data should be referred to KSM, Maccabi Healthcare Services Research and Innovation Center.

## Code Availability

The source code is available at https://github.com/yakiSaciuk/Estimating-Penic-AMR-using-IV. The repository contains R scripts for estimating risk differences (RD) using two-stage least squares (2SLS) instrumental variable analysis, deriving confidence intervals via bootstrap sampling, while applying inverse probability of censoring weights (IPCW). The code is provided under the MIT License.

## Acknowledgements

This work was supported by the Israel Science Foundation (ISF 1286/21) - UO

## Author Contributions Statement

Conceptualization: YS, UO

Methodology: YS, DN, UO

Investigation: YS, MC, UO

Visualization: YS

Funding acquisition: UO

Supervision: UO

Writing – original draft: YS, MC

Writing – review & editing: DN, MC, UO

