## Supplementary Information for "Penicillin Allergy as an Instrumental Variable for Estimating Antibiotic Effects on Resistance"

### Table S1: First Stage Regression Coefficients for Instrumental Variable (IV) Estimation

| | Amoxicillin/ clavulanic acid | Ampicillin | Piperacillin/Tazobactam | Gentamicin |
|---|---|---|---|---|
| IV *Coeff. (S.E.)*(1) | 0.21(0.01)*** | 0.24(0.01)*** | 0.23(0.01)*** | 0.23(0.01)*** |
| Kleibergen-Paap F-statistics (2) | 751.5*** | 1,347.2*** | 1,588.5**** | 1,649.8*** |

*** p-value < 0.001, **p -value < 0.01,
* p-value < 0.05

(1) Adjusted for sex, age, ses, social sector, comorbidities (ckd, cardio-vascular disease, diabetes mellitus, hypertension and immunosuppression), calendar time, previous urine culture tests count, and previous antibiotic uses count. Standard errors are clustered at the individual level to account for within-individual correlation in the data. The p-value for the instrument coefficient in all first-stage regressions was less than 0.01.

(2) Kleibergen-Paap F-statistics were computed with standard errors clustered at the individual level, with all associated p-values remaining below 0.01 across all regressions.

### Table S2: Instrumental Variable (IV)-Estimated Risk Differences of Antimicrobial Resistance (AMR) Rates: A Comparison between Exposure to Penicillins and Other Antibiotics Across Varying Time Points Post-Exposure – Without Inverse Probability Censoring Weights (IPCW) Weighting

| | | | | 95% C.I. (1) | | |
|---|---|---|---|---|---|---|
| **AMR Antibiotic** | **Follow up length** | **Number of AMR Tests** | **RD IV (1)** | **Lower Bound (2.5%)** | **Upper Bound (97.5%)** | **NNH** |
| Amoxicillin/clavulanic acid | Up to: 90 days | 7,502 | 30.0% | 19.7% | 42.4% | 3.3 |
| | Up to: 180 days | 12,393 | 21.7% | 13.5% | 29.5% | 4.6 |
| | Up to: 270 days | 16,027 | 16.5% | 10.5% | 23.2% | 6.1 |
| | Up to: 365 days | 18,797 | 15.2% | 9.8% | 21.4% | 6.6 |
| Ampicillin | Up to: 90 days | 13,776 | 21.2% | 11.3% | 33.8% | 4.7 |
| | Up to: 180 days | 21,828 | 16.6% | 8.7% | 26.1% | 6.0 |

| | | | | | | |
|---|---|---|---|---|---|---|
| | Up to: 270 days | 27,587 | 12.3% | 4.5% | 19.9% | 8.1 |
| | Up to: 365 days | 32,006 | 11.8% | 5.5% | 20.3% | 8.5 |
| Piperacillin/Tazobactam | Up to: 90 days | 15,203 | 2.5% | 1.1% | 3.7% | 40.8 |
| | Up to: 180 days | 24,639 | 1.8% | 0.9% | 2.6% | 55.6 |
| | Up to: 270 days | 31,730 | 1.3% | 0.3% | 2.4% | 75.2 |
| | Up to: 365 days | 37,295 | 1.0% | 0.3% | 1.9% | 98.0 |
| Gentamicin (Negative Control) | Up to: 90 days | 17,346 | -0.9% | -5.8% | 3.9% | -114.9* |
| | Up to: 180 days | 27,824 | -1.7% | -6.4% | 3.0% | -60.2* |
| | Up to: 270 days | 35,447 | -1.5% | -5.0% | 2.2% | -67.6* |
| | Up to: 365 days | 41,317 | 0.2% | -2.8% | 2.7% | 555.6 |

(1) Adjusted for sex, age, ses, social sector, comorbidities (ckd, cardio-vascular disease, diabetes mellitus, hypertension and immunosuppression), calendar time, previous urine culture tests count, and previous antibiotic uses count.

* The negative NNH is interpreted as the absolute value of the NNH when switching the penicillins to be the comparator group.

Table S3: Instrumental Variable (IV)-Estimated Risk Differences of Antimicrobial Resistance (AMR) Rates: A Comparison between Exposure to Penicillins and Other Antibiotics Across Varying Time Points Post-Exposure – Matched Analysis

| | | | | 95% C.I. (1)(2) | | |
|---|---|---|---|---|---|---|
| **AMR Antibiotic** | **Follow up length** | **Number of AMR Tests** | **RD IV (1)(2)** | **Lower Bound (2.5%)** | **Upper Bound (97.5%)** | **NNH** |
| Amoxicillin/clavulanic acid | Up to: 90 days | 7,502 | 15.1% | 4.7% | 24.9% | 6.6 |
| | Up to: 180 days | 12,393 | 14.0% | 7.0% | 20.4% | 7.1 |
| | Up to: 270 days | 16,027 | 9.6% | 4.3% | 14.7% | 10.4 |
| | Up to: 365 days | 18,797 | 9.0% | 4.0% | 13.7% | 11.1 |
| Ampicillin | Up to: 90 days | 13,776 | 27.7% | 17.2% | 38.3% | 3.6 |
| | Up to: 180 days | 21,828 | 22.0% | 14.3% | 30.1% | 4.5 |
| | Up to: 270 days | 27,587 | 17.3% | 10.3% | 24.4% | 5.8 |
| | Up to: 365 days | 32,006 | 15.5% | 9.4% | 21.5% | 6.5 |
| Piperacillin/Tazobactam | Up to: 90 days | 15,203 | 2.6% | 1.6% | 3.4% | 39.2 |
| | Up to: 180 days | 24,639 | 1.6% | 0.8% | 2.3% | 61.7 |
| | Up to: 270 days | 31,730 | 1.2% | 0.5% | 1.8% | 85.5 |
| | Up to: 365 days | 37,295 | 1.1% | 0.5% | 1.6% | 92.6 |
| Gentamicin (Negative Control) | Up to: 90 days | 17,346 | -4.5% | -9.7% | 0.3% | -22.4* |
| | Up to: 180 days | 27,824 | -3.5% | -7.5% | 0.1% | -28.3* |
| | Up to: 270 days | 35,447 | -2.1% | -5.1% | 0.7% | -46.7* |

| | | | | | | |
|---|---|---|---|---|---|---|
| | Up to: 365 days | 41,317 | -0.9% | -3.6% | 1.7% | -116.3* |

(1) Weighted with inverse probability of censoring (IPCW) weights

(2) Adjusted and matched with full matching for sex, age, ses, social sector, comorbidities (ckd, cardio-vascular disease, diabetes mellitus, hypertension and immunosuppression), calendar time, previous urine culture tests count, and previous antibiotic uses count. The first-stage regression used a probit model, with fitted probabilities used to estimate the effect in the second stage.

* The negative NNH is interpreted as the absolute value of the NNH when switching the penicillins to be the comparator group.

Table S4: Instrumental Variable (IV)-Estimated Risk Differences of Antimicrobial Resistance (AMR) Rates: A Comparison between Exposure to Penicillins and Other Antibiotics Across Varying Time Points Post-Exposure, Limited to Amoxicillin, Amoxicillin without Other Penicillins, and Amoxicillin with Other Penicillins

| | | | | | 95% C.I. (1)(2) | | |
|---|---|---|---|---|---|---|---|
| **Penicillin** | **AMR Antibiotic** | **Follow up length** | **Number of AMR Tests** | **RD IV (1)(2)** | **Lower Bound (2.5%)** | **Upper Bound (97.5%)** | **NNH** |
| Amoxicillin/ clavulanic acid | Amoxicillin/clavulanic acid | Up to: 90 days | 6,455 | 31.2% | 21.5% | 41.3% | 3.2 |
| | | Up to: 180 days | 10,362 | 22.4% | 14.3% | 30.4% | 4.5 |
| | | Up to: 270 days | 13,226 | 17.4% | 10.8% | 23.8% | 5.7 |
| | | Up to: 365 days | 15,356 | 15.7% | 10.1% | 21.5% | 6.4 |
| | Ampicillin | Up to: 90 days | 11,238 | 28.5% | 15.6% | 41.8% | 3.5 |
| | | Up to: 180 days | 17,510 | 22.6% | 13.0% | 33.2% | 4.4 |
| | | Up to: 270 days | 21,918 | 20.2% | 11.3% | 28.6% | 5.0 |
| | | Up to: 365 days | 25,253 | 16.9% | 9.7% | 25.0% | 5.9 |
| | Piperacillin/ Tazobactam | Up to: 90 days | 12,655 | 4.1% | 2.6% | 5.6% | 24.3 |
| | | Up to: 180 days | 20,111 | 2.9% | 1.9% | 3.8% | 34.7 |
| | | Up to: 270 days | 25,611 | 1.6% | 0.5% | 2.5% | 64.5 |
| | | Up to: 365 days | 29,865 | 1.0% | 0.1% | 1.8% | 105.3 |
| | Gentamicin | Up to: 90 days | 14,356 | -0.2% | -6.5% | 5.8% | -500.0* |
| | | Up to: 180 days | 22,629 | 0.7% | -3.6% | 4.6% | 153.8 |

| | | | | | | | |
|---|---|---|---|---|---|---|---|
| | | Up to: 270 days | 28,524 | -0.2% | -3.8% | 3.3% | -454.5* |
| | | Up to: 365 days | 33,006 | 1.1% | -2.0% | 4.2% | 88.5 |
| Amoxicillin | Amoxicillin/clavulanic acid | Up to: 90 days | 6,459 | 24.7% | 14.0% | 35.9% | 4.0 |
| | | Up to: 180 days | 10,540 | 20.4% | 13.0% | 27.7% | 4.9 |
| | | Up to: 270 days | 13,464 | 15.6% | 9.3% | 21.7% | 6.4 |
| | | Up to: 365 days | 15,684 | 15.0% | 9.6% | 20.4% | 6.7 |
| | Ampicillin | Up to: 90 days | 11,731 | 30.3% | 18.5% | 43.0% | 3.3 |
| | | Up to: 180 days | 18,396 | 23.6% | 14.6% | 32.6% | 4.2 |
| | | Up to: 270 days | 23,049 | 20.9% | 12.5% | 28.7% | 4.8 |
| | | Up to: 365 days | 26,585 | 19.5% | 12.3% | 26.8% | 5.1 |
| | Piperacillin/ Tazobactam | Up to: 90 days | 13,106 | 2.3% | 1.3% | 3.3% | 43.7 |
| | | Up to: 180 days | 20,960 | 1.7% | 0.8% | 2.4% | 59.9 |
| | | Up to: 270 days | 26,710 | 0.9% | 0.0% | 1.6% | 116.3 |
| | | Up to: 365 days | 31,197 | 0.6% | -0.2% | 1.3% | 161.3 |
| | Gentamicin | Up to: 90 days | 14,889 | -3.1% | -9.3% | 2.4% | -31.9* |
| | | Up to: 180 days | 23,591 | -2.7% | -6.9% | 1.2% | -36.5* |
| | | Up to: 270 days | 29,772 | -2.4% | -5.8% | 1.0% | -41.5* |
| | | Up to: 365 days | 34,494 | -0.4% | -3.3% | 2.6% | -270.3* |
| Amoxicillin and Penicillin | Amoxicillin/clavulanic acid | Up to: 90 days | 6,705 | 20.5% | 11.3% | 30.7% | 4.9 |
| | | Up to: 180 days | 10,998 | 17.7% | 11.3% | 24.5% | 5.7 |
| | | Up to: 270 days | 14,103 | 13.7% | 7.5% | 19.2% | 7.3 |
| | | Up to: 365 days | 16,468 | 13.6% | 8.6% | 18.2% | 7.4 |
| | Ampicillin | Up to: 90 days | 12,219 | 28.2% | 17.5% | 39.8% | 3.5 |
| | | Up to: 180 days | 19,236 | 22.1% | 13.5% | 30.9% | 4.5 |
| | | Up to: 270 days | 24,161 | 19.0% | 12.0% | 26.1% | 5.3 |
| | | Up to: 365 days | 27,943 | 17.7% | 11.3% | 24.4% | 5.6 |
| | Piperacillin/ Tazobactam | Up to: 90 days | 13,612 | 1.7% | 0.5% | 2.6% | 59.5 |
| | | Up to: 180 days | 21,871 | 1.2% | 0.5% | 1.9% | 81.3 |
| | | Up to: 270 days | 27,953 | 0.6% | -0.2% | 1.3% | 163.9 |
| | | Up to: 365 days | 32,735 | 0.5% | -0.3% | 1.2% | 208.3 |
| | Gentamicin | Up to: 90 days | 15,489 | -3.2% | -8.5% | 1.7% | -31.7* |
| | | Up to: 180 days | 24,640 | -2.8% | -6.5% | 0.6% | -35.8* |
| | | Up to: 270 days | 31,179 | -2.3% | -5.4% | 0.6% | -43.5* |
| | | Up to: 365 days | 36,214 | -0.6% | -3.4% | 2.0% | -163.9* |

(1) Weighted with inverse probability of censoring (IPCW) weights

(2) Adjusted for sex, age, ses, social sector, comorbidities (ckd, cardio-vascular disease, diabetes mellitus, hypertension and immunosuppression), calendar time, previous urine culture tests count and previous antibiotic uses count

* The negative NNH is interpreted as the absolute value of the NNH when switching the penicillins to be the comparator group.

Trimethoprim/sulfamethoxazole

Table S5: Instrumental Variable (IV)-Estimated Risk Differences Across Antimicrobial Resistance (AMR) Test Strata: Pairwise Comparisons of penicillin vs. Other Antibiotic Groups Within 180 Days Post-Exposure

| AMR Antibiotic | Paired Exposure Antibiotic | Number of AMR Tests | RD IV (1)(2) | 95% C.I. (1)(2) Lower Bound (2.5%) | 95% C.I. (1)(2) Upper Bound (97.5%) | NNH |
|---|---|---|---|---|---|---|
| Amoxicillin/ clavulanic acid | **All Groups** | 12,393 | 14.9% | 9.9% | 20.2% | 6.7 |
| | Cephalosporins | 6,420 | 16.5% | -0.5% | 33.2% | 6.1 |
| | Macrolides | 4,532 | 10.3% | 4.9% | 15.3% | 9.7 |
| | Tetracyclines | 3,695 | 21.3% | -7.0% | 54.0% | 4.7 |
| | Trimethoprim/sulfamethoxazole | 3,770 | 73.2% | -44.4% | 364.8% | 1.4 |
| | Fosfomycin Trometamol | 5,181 | 32.2% | -3.6% | 81.9% | 3.1 |
| | Nitrofurantoin | 4,402 | 50.7% | -4.4% | 135.8% | 2.0 |
| Ampicillin | **All Groups** | 21,828 | 18.2% | 11.9% | 24.6% | 5.5 |
| | Cephalosporins | 11,670 | 9.9% | -6.1% | 25.8% | 10.1 |
| | Macrolides | 8,683 | 21.5% | 14.5% | 28.1% | 4.7 |
| | Tetracyclines | 7,360 | 32.8% | 3.4% | 64.8% | 3.0 |
| | Trimethoprim/sulfamethoxazole | 7,582 | 116.9% | -19.9% | 436.8% | 0.9 |

| | | | | | | |
|---|---|---|---|---|---|---|
| | Fosfomycin Trometamol | 9,506 | 36.7% | -4.0% | 82.7% | 2.7 |
| | Nitrofurantoin | 8,399 | 64.6% | 6.5% | 141.9% | 1.5 |
| Piperacillin/ Tazobactam | **All Groups** | 24,639 | 1.6% | 0.9% | 2.2% | 61.7 |
| | Cephalosporins | 12,791 | 1.7% | -0.6% | 3.5% | 58.1 |
| | Macrolides | 9,390 | 1.4% | 0.5% | 2.2% | 69.9 |
| | Tetracyclines | 7,799 | 4.1% | -0.7% | 8.0% | 24.6 |
| | Trimethoprim/sulfamethoxazole | 8,033 | 11.0% | -16.4% | 52.4% | 9.1 |
| | Fosfomycin Trometamol | 10,627 | 3.8% | -2.6% | 9.3% | 26.5 |
| | Nitrofurantoin | 9,127 | 5.4% | -3.4% | 13.0% | 18.6 |
| Gentamicin | **All Groups** | 27,824 | -1.5% | -4.5% | 1.3% | -66.2* |
| | Cephalosporins | 14,591 | -6.6% | -16.4% | 1.1% | -15.2* |
| | Tetracyclines | 8,954 | -9.5% | -32.2% | 7.7% | -10.5* |
| | Trimethoprim/sulfamethoxazole | 9,225 | -54.0% | -257.3% | 20.5% | -1.9* |
| | Fosfomycin Trometamol | 11,881 | -15.4% | -40.1% | 3.9% | -6.5* |
| | Nitrofurantoin | 10,351 | -22.8% | -73.7% | 10.5% | -4.4* |

(1) Weighted with inverse probability of censoring (IPCW) weights

(2) Adjusted for sex, age, ses, social sector, comorbidities (ckd, cardio-vascular disease, diabetes mellitus, hypertension and immunosuppression), calendar time, previous urine culture tests count and previous antibiotic uses count

* The negative NNH is interpreted as the absolute value of the NNH when switching the penicillins to be the comparator group.

Figure S1: Resistance Trends of Selected Antibiotics Against *E. coli* in Urine Cultures 2013-2022, Maccabi Healthcare Services (MHS)

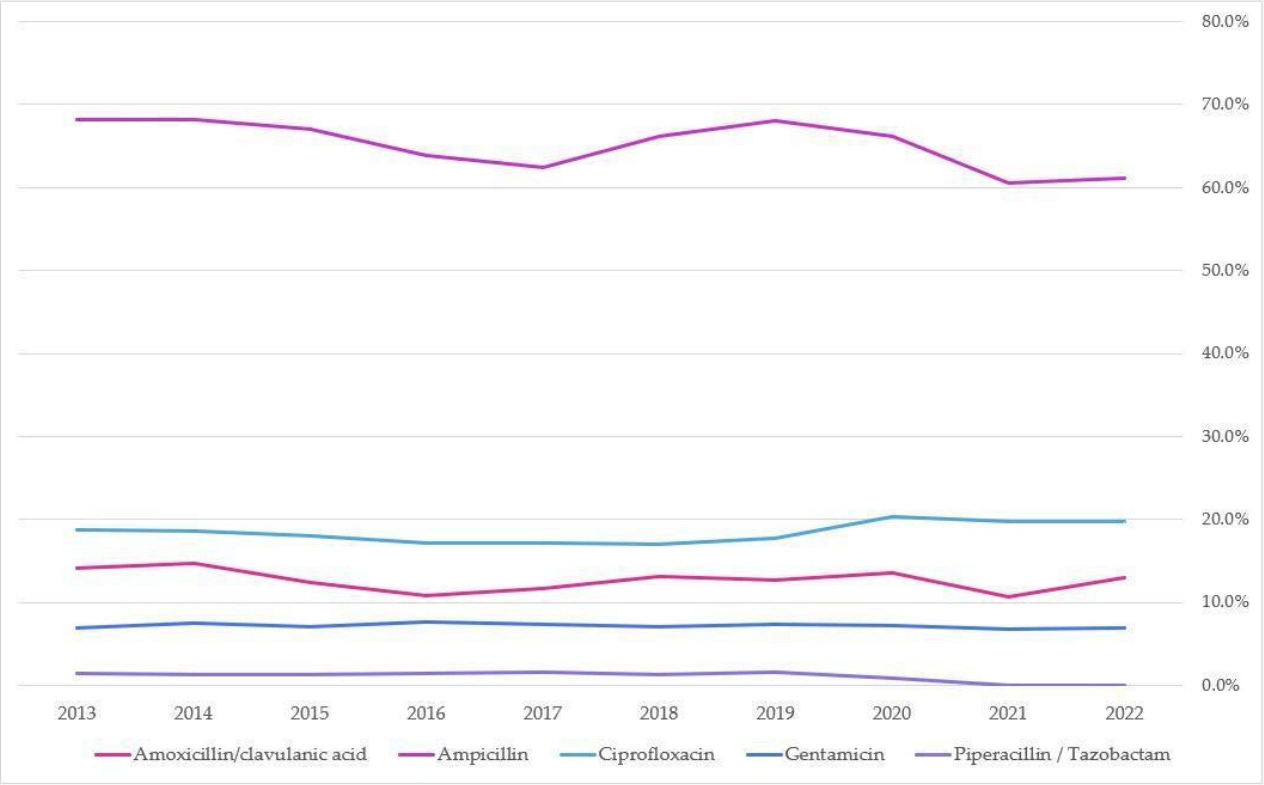

Figure S2: Assumed Causal Model Represented as a Directed Acyclic Graph (DAG) with Named Covariates

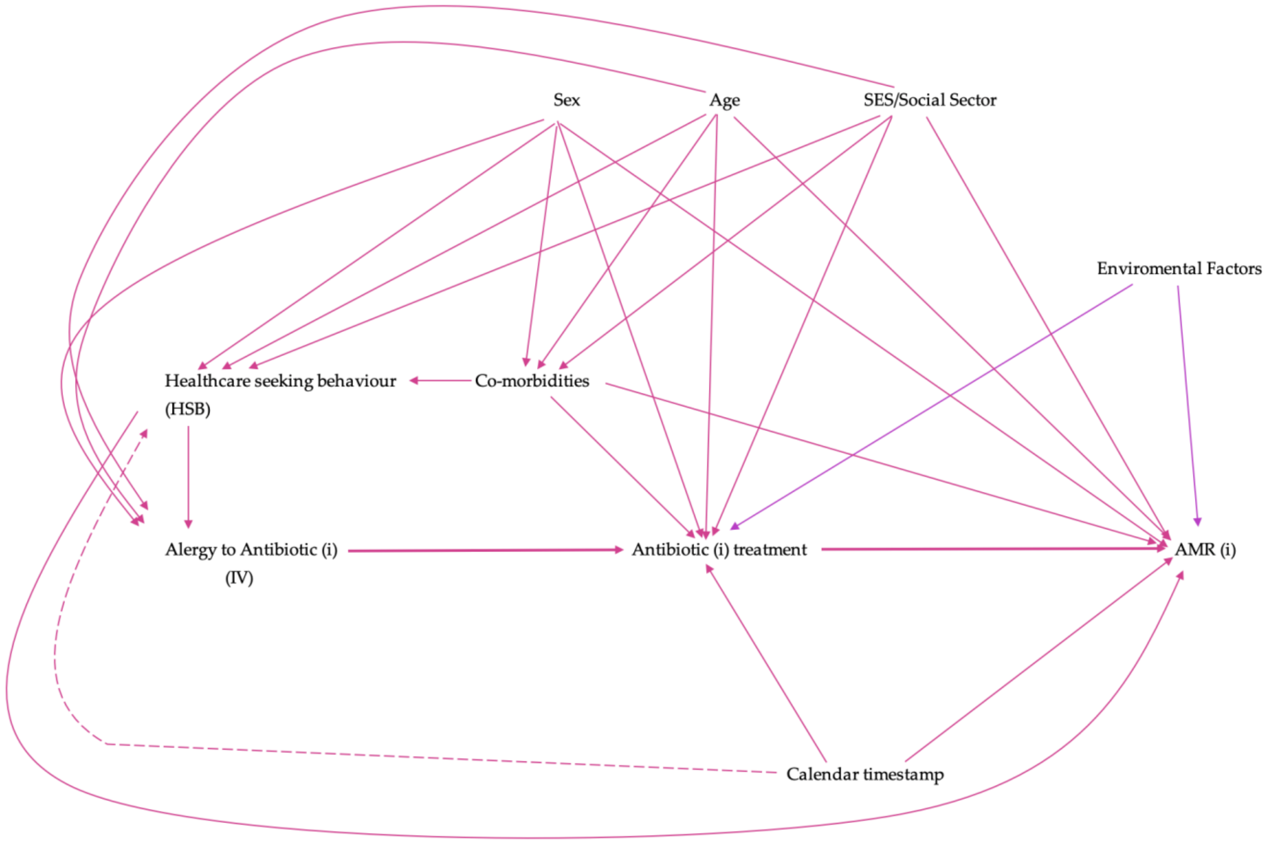


Table S6: Probability of being uncensored in the primary study population.

| | Status | N | Min. | 1st Qu. | Median | Mean | 3rd Qu. | Max. |
|---|---|---|---|---|---|---|---|---|
| Amoxicillin/ clavulanic acid | uncensored | 18,797 | 0.05% | 0.56% | 1.13% | 1.37% | 1.86% | 11.10% |
| | censored | 3,087,294 | 0.00% | 0.08% | 0.48% | 0.60% | 0.71% | 11.13% |
| Ampicillin | uncensored | 32,006 | 0.09% | 0.97% | 1.77% | 2.24% | 2.91% | 18.66% |
| | censored | 3,074,085 | 0.00% | 0.15% | 0.84% | 1.02% | 1.28% | 20.35% |
| Piperacillin/ Tazobactam | uncensored | 37,295 | 0.09% | 1.12% | 2.13% | 2.71% | 3.61% | 23.87% |
| | censored | 3,068,796 | 0.00% | 0.17% | 0.94% | 1.18% | 1.43% | 24.99% |
| Gentamicin | uncensored | 41,317 | 0.11% | 1.24% | 2.36% | 2.95% | 3.86% | 23.62% |
| | censored | 3,064,774 | 0.00% | 0.19% | 1.06% | 1.31% | 1.59% | 25.41% |
| Ciprofloxacin | uncensored | 37,516 | 0.10% | 1.12% | 2.14% | 2.65% | 3.47% | 21.95% |
| | censored | 3,068,575 | 0.00% | 0.18% | 0.96% | 1.19% | 1.45% | 23.52% |
